# Out-of-sequence placement of deceased donor kidneys is exacerbating inequities in the United States

**DOI:** 10.1101/2025.02.27.25322934

**Authors:** Sumit Mohan, Miko E. Yu, Joel T. Adler, Lindsey M. Maclay, David C. Cron, Jesse Schold, Syed Ali Husain

## Abstract

**Background:** Deceased donor kidney allocation in the United States follows an objective algorithm that is designed to balance equity and utility. Organ procurement organizations (OPOs) are increasingly using out-of-sequence allocation of deceased donor kidneys (DDKs) to improve organ utilization rates. We investigated trends in the prevalence of out-of-sequence allocation in DDKs, and the association with successful organ placement, and its impact on equity and organ utilization.

**Methods:** Using 2020-2023 transplant data from the Scientific Registry of Transplant Recipients and organ offer data from the potential transplant recipient dataset, we identified all organ offers for DDKs, including those placed out-of-sequence, along with donor and recipient characteristics. We obtained the prevalence of out-of-sequence allocation and placement in DDKs and assessed temporal trends across organ quality as measured by the kidney donor profile index (KDPI). Lorenz curves and Gini coefficients measured inequality for out-of-sequence transplantation at the transplant center and OPO levels.

**Results:** From 2020 to 2023, out-of-sequence placement of DDKs increased from 340 kidneys in 2020 (2.3%) to 2,931 in 2023 (15.8%). Out-of-sequence placements now account for 1-in-7 DDK transplants and include kidneys across the organ quality spectrum, with 41% of these occurring with KDPI<50% (i.e., the highest quality) kidneys. By 2023, 91% of OPOs were using out-of-sequence allocation, reflected in the Gini coefficient decreasing from 0.89 to 0.47 with no appreciable impact on utilization rates.

**Conclusions:** There has been a sharp increase in out-of-sequence allocation of DDKs including high-quality organs. Most of these kidneys are being placed at a small number of transplant centers, creating preferential access to transplantation and exacerbating inequities in access to transplantation.

**Key Points:**

1. Out-of-sequence transplantation of deceased donor kidneys has increased dramatically from 2020 to 2023 without regard for organ quality.
2. A limited number of centers are responsible for a third of all out-of-sequence transplants, exacerbating inequality in access to transplants.
3. There has been no improvement in the utilization of deceased donor kidneys despite the increase of out-of-sequence placements.

## Introduction

Kidney transplantation is the ideal treatment for patients with end-stage kidney disease, but there is a shortage of organs donated for transplant. In the United States (US), most transplants utilize organs from deceased donors, which are allocated using a points-based policy that ranks potential candidates on priority for an organ when it becomes available.[1–4] Prioritization for deceased donor kidneys is determined primarily by allocation time but is also influenced by other factors such as the distance between the organ and recipient, allosensitization, and prioritization of select groups, including children, former living donors, and highly sensitized patients.[3 4]

Despite the long waiting times for a kidney transplant throughout the country, 99.7% of organ offers to waitlisted transplant candidates are declined, typically because a candidate or transplant center believes it is better to wait for a future, higher-quality organ offer.[5] When an offer for a deceased donor kidney is declined, the organ is sequentially prioritized for the next individual on the waitlist that is generated specifically for that organ using a predefined objective, algorithmic allocation system designed to balance equity and utility.[5 6] This organ specific prioritization of patients is referred to as the “match run”. Changes in the allocation system introduced in 2021 with a goal of reducing geographic disparities have adversely impacted allocation efficiency. This change in the allocation system appears to be driving sharp increases in a greater number of centers declining organs which in turn has resulted in a rising rate of deceased donor kidney being discarded rather than being successfully transplanted, particularly for less-than-ideal quality deceased donor organs.[7–11]

Organ procurement organizations (OPOs) are responsible for organ recovery, facilitating the logistical process of offering organs to transplant centers, identifying the center that eventually accepts the organ, and then transporting the organ to that center. Increased regulatory oversight of OPO performance, including their successful placement of available deceased donor organs, has also been recently implemented.[12 13] The current inefficiencies of the system have contributed to the frequent decline of organ offers and the subsequent risk of non-utilization of less-than-ideal organs. OPOs have the ability to circumvent the stepwise allocation process and ignore the sequence in the match run – an exception that was rarely invoked in prior years. More recently, however, OPOs have increasingly opted to offer a given organ directly to a transplant center that they believe would be willing to accept that organ even if that means overlooking patients and centers that have a higher priority for the available deceased donor kidney on the match run.[14 15] Because these “out-of-sequence” allocations violate the enshrined allocation policy, each instance requires an explanation of the extenuating circumstances that led to the need to deviate from the allocation algorithm and is reviewed for appropriateness by the Organ Procurement and Transplantation Network (OPTN).[14 16]

While these out-of-sequence placements have historically occurred in approximately 1% of deceased donor kidneys as a salvage allocation strategy, we aim to investigate trends in the prevalence of out-of-sequence allocation in deceased donor kidneys, examine the association with successful organ placement, and understand its impact on equity in access to transplantation.[14]

## Methods

We performed a retrospective cohort study using U.S. transplant registry data and potential recipient data. This study used data from the Scientific Registry of Transplant Recipients (SRTR). The SRTR data system includes data on all donors, wait-listed candidates, and transplant recipients in the US, submitted by the members of the Organ Procurement and Transplantation Network (OPTN). The Health Resources and Services Administration (HRSA), U.S. Department of Health and Human Services, provides oversight to the activities of the OPTN and SRTR contractors. The data reported here have been supplied by the Hennepin Healthcare Research Institute (HHRI) as the contractor for the Scientific Registry of Transplant Recipients (SRTR). The interpretation and reporting of these data are the responsibility of the author(s) and in no way should be seen as an official policy of or interpretation by the SRTR or the U.S. Government. This study used potential transplant recipient (PTR) match run data available through 2023 and the standard analysis files from the SRTR dated December 2024. The Columbia University Irving Medical Center institutional review board approved this study. All research activities were consistent with the principles of the Declaration of Istanbul.

We identified all match runs for deceased donor kidneys in the US (n=60,166) from 2020 through 2023 (Supplemental Figure 1). Match runs from Puerto Rico (n=366) and bypassed offers except those with out-of-sequence codes (n= 590 match runs) were excluded. Out-of-sequence offers were defined as offers with primary or secondary refusal codes 861: Operational OPO, 862: Donor medical urgency, or 863: Offer not made due to expedited placement attempt. Organ offers that followed allocation policy are referred to as “in-sequence” allocations.

We evaluated OPO characteristics, including discard rate, calculated using the number of kidneys recovered, the number of kidneys recovered for transplantation but not transplanted, and the proportion of kidneys offered out-of-sequence among all recovered kidneys. The kidney donor profile index (KDPI), an indicator of organ quality that is used as part of the allocation algorithm, was calculated using the 2022 scaling factor.[4] Since higher KDPI scores suggest shorter graft survival, the current OPTN policy requires patients to be educated about their options. Patients must proactively opt-in to receive offers for “high KDPI” (KDPI>85%) kidneys with written consent before they can be considered for organs in this category. Consequently, the match run for kidneys with a KDPI>85% only includes patients who have consented to accept these kidneys. We examined overall and temporal changes in out-of-sequence rates and discard rates among all recovered kidneys and high KDPI kidneys. We also evaluated correlations between OPO discard rates and out-of-sequence organ offers for these strata.

We calculated the proportion of out-of-sequence organs as a fraction of all deceased donor kidney transplants performed at centers over the duration of the study. We also looked at the proportion across the different KDPI deciles. Additionally, we studied changes over time at both the transplant center and OPO levels.

We constructed Lorenz curves and calculated the Gini coefficients as a measure of inequality in the distribution of both in-sequence and out-of-sequence sequence deceased donor kidney transplantation at both the transplant center and OPO levels. The Lorenz curve and Gini coefficient were originally developed as a measure of wealth concentration[17] but have been applied to measure health disparities. For instance, in Figure 2, the cumulative population of OPOs is represented on the X-axis and the cumulative proportion of “wealth” (i.e., transplants) on the Y-axis.[18] Gini coefficients range from 0-1, with 1 representing maximal inequality and values above 0.4 generally considered to reflect high levels of inequality.[19] We compared temporal trends and changes over time, including for high KDPI deceased donor kidneys.

We compared the characteristics of recipients who received an out-of-sequence transplant with those who received an in-sequence transplant. These characteristics included age at transplant, race, ethnicity, gender, diabetes history, education, primary payer at transplant, cold ischemia time (CIT), median KDPI, preemptive transplant status, and dialysis time if not preemptively transplanted.

We compared center and recipient characteristics using chi-squared and Wilcoxon rank-sum tests. A two-sided alpha of 0.05 determined statistical significance. Analyses were performed using STATA/MP 17.0 (StataCorp, College Station, TX).

## Results

### Rapid increase in out-of-sequence transplants

From 2020 to 2023, the number of deceased donor kidney transplants performed in the US increased by 24%. During the same period, out-of-sequence placement of deceased donor kidneys increased disproportionately from 340 kidneys in 2020 (2.3%) to 2,931 in 2023 (15.8%) – an 8-fold increase in the proportion of deceased donor kidney transplants performed out-of-sequence (Figure 1). In 2023, 15.8% of deceased donor kidneys – nearly 1 in 7 deceased donor kidney transplants were placed with candidates out-of-sequence. The median KDPI of kidneys placed out-of-sequence increased over time from 52% in 2020 to 57% in 2023.

**Figure 1.**
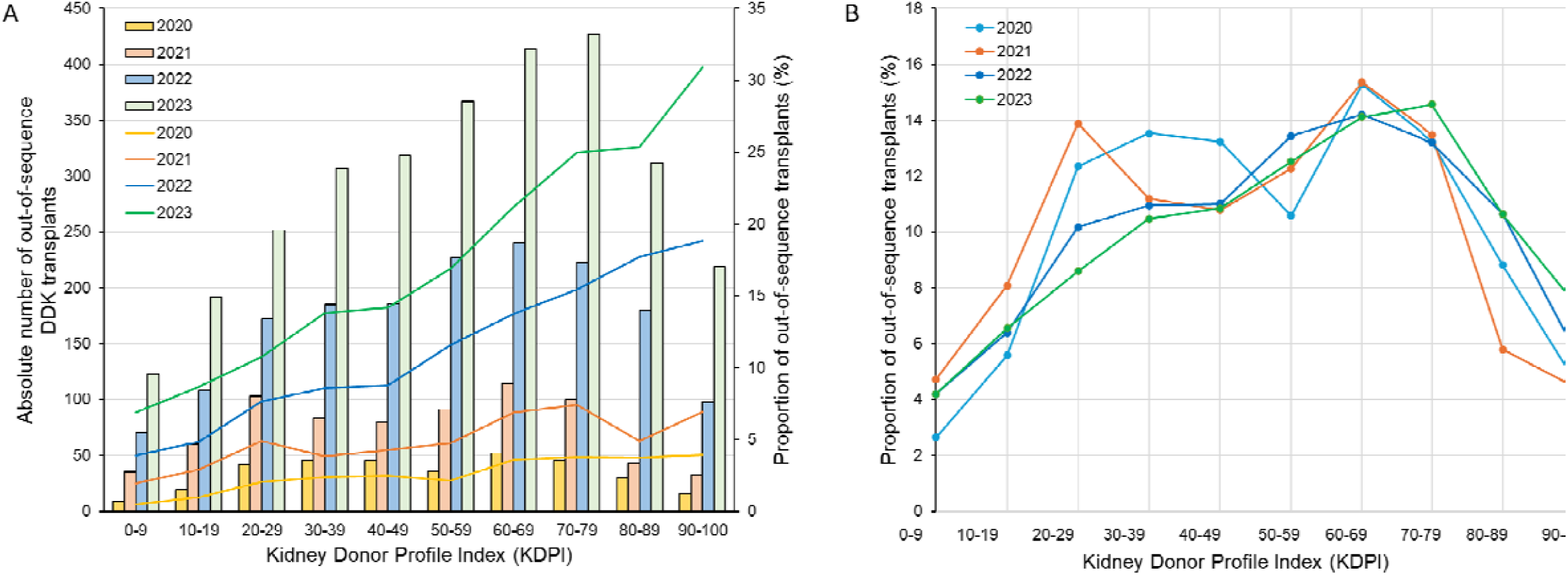
Distribution of out-of-sequence deceased donor kidneys within KDPI deciles, 2020-2023. Panel A shows the absolute number of out-of-sequence kidneys transplanted and the proportion of all deceased donor kidney transplants that were out-of-sequence in each of the 4 years. Panel B shows the proportion of out-of-sequence deceased donor kidney transplants within each KDPI decile each year.

When stratifying deceased donor kidney transplants by KDPI decile, we noted an increasing proportion of kidneys placed out-of-sequence in higher deciles, along with sharply increasing proportions of deceased donor kidneys placed out-of-sequence across years within each decile (Figure 1) and a greater rate of increase in out-of-sequence placements with increasing KDPI. By 2023, this trend resulted in the proportion of out-of-sequence deceased donor kidney transplants rising to 6.9% in the lowest decile and 30.9% for the highest decile (Supplemental Table 1). Between 2020 and 2023, we noted a 5- to 10-fold increase in the proportion of kidneys transplanted as out-of-sequence kidneys within each decile. Over a third (40.7%) of all out-of-sequence transplants occurred with a KDPI <50% deceased donor kidney, including 1,192 kidneys in 2023 alone and representing 11% of all KDPI <50% transplants. The distribution of KDPI deciles among out-of-sequence transplants stayed relatively constant across years (Figure 1).

### Changing OPO organ offer practices

In 2020, even with the ongoing pandemic, very few of the 57 OPOs facilitated the majority of the out-of-sequence transplants performed, while 36 OPOs did not place any kidneys out-of-sequence. This was reflected by a high Gini coefficient of 0.89 for out-of-sequence transplants (vs. 0.31 for in-sequence transplants) at the level of the OPOs (Figure 2). A total of 340 deceased donor kidneys were offered out-of-sequence, which included 2 OPOs offering more than 10% of deceased donor kidneys out-of-sequence – with one of these OPOs offering 3 times the number of organs out-of-sequence as the other.

**Figure 2.**
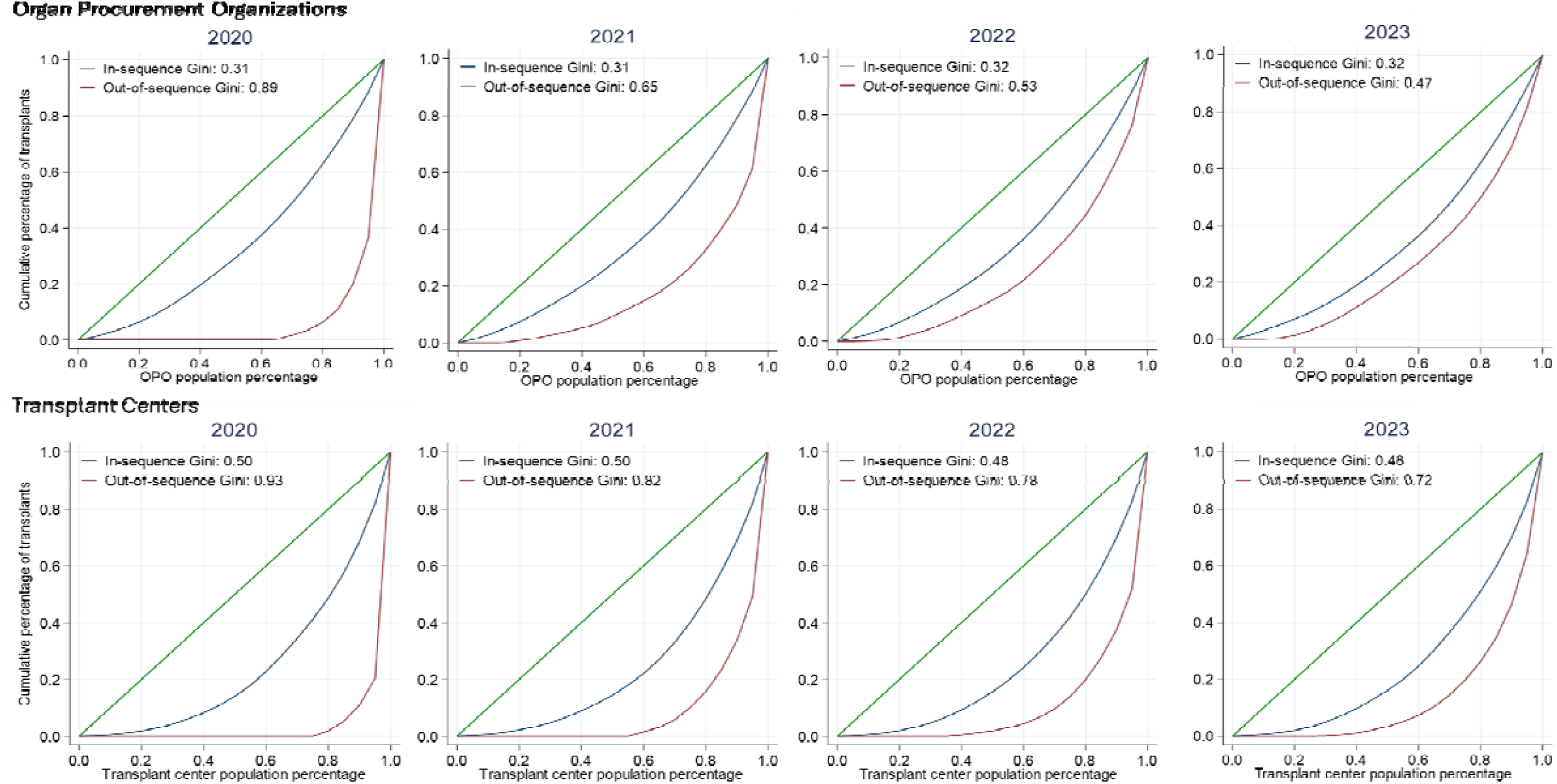
Lorenz curves for the cumulative percentage of transplants for all OPOs and transplant centers, stratified by kidneys that were allocated either in-sequence or out-of-sequence for each year in the study period. The Gini coefficient ranges from 0, representing perfect equality and depicted as the green diagonal line, to 1, representing perfect inequality.

By 2023, the Gini coefficient for out-of-sequence transplants had decreased to 0.47 (vs. 0.32 for in-sequence transplants), reflecting the broad and rapid uptake of the practice of out-of-sequence placements by most OPOs (Figure 2). Only 5 of the 55 OPOs did not offer any deceased donor kidneys out-of-sequence in 2023. Further, there was increased utilization of out-of-sequence placement for more deceased donor kidneys by individual OPOs, as evidenced by the 11 OPOs that offered more than a quarter of their deceased donor kidneys out-of-sequence, including one OPO that offered 55% of their deceased donor kidneys out-of-sequence.

Between 2020 and 2023, only 4 OPOs experienced small absolute decreases in their discard rate and among these, 3 centers had an absolute increase (5-15%) in their proportion of deceased donor kidneys offered out-of-sequence (Figure 3). At the higher end of the KDPI scale, small absolute decreases in OPO discard rates also occurred (r=-0.42, p=0.001). Only one OPO appears to have substantially lowered the proportion of organs offered out-of-sequence between 2020 and 2023. During the study period, 3 unique OPO-transplant center pairs accounted for 100 or more out-of-sequence transplants each; 2 of these 3 pairings included the same OPO. In comparison, the median number of out-of-sequence transplants among the remaining 1,286 unique OPO-transplant centers pairs was only 2.

**Figure 3.**
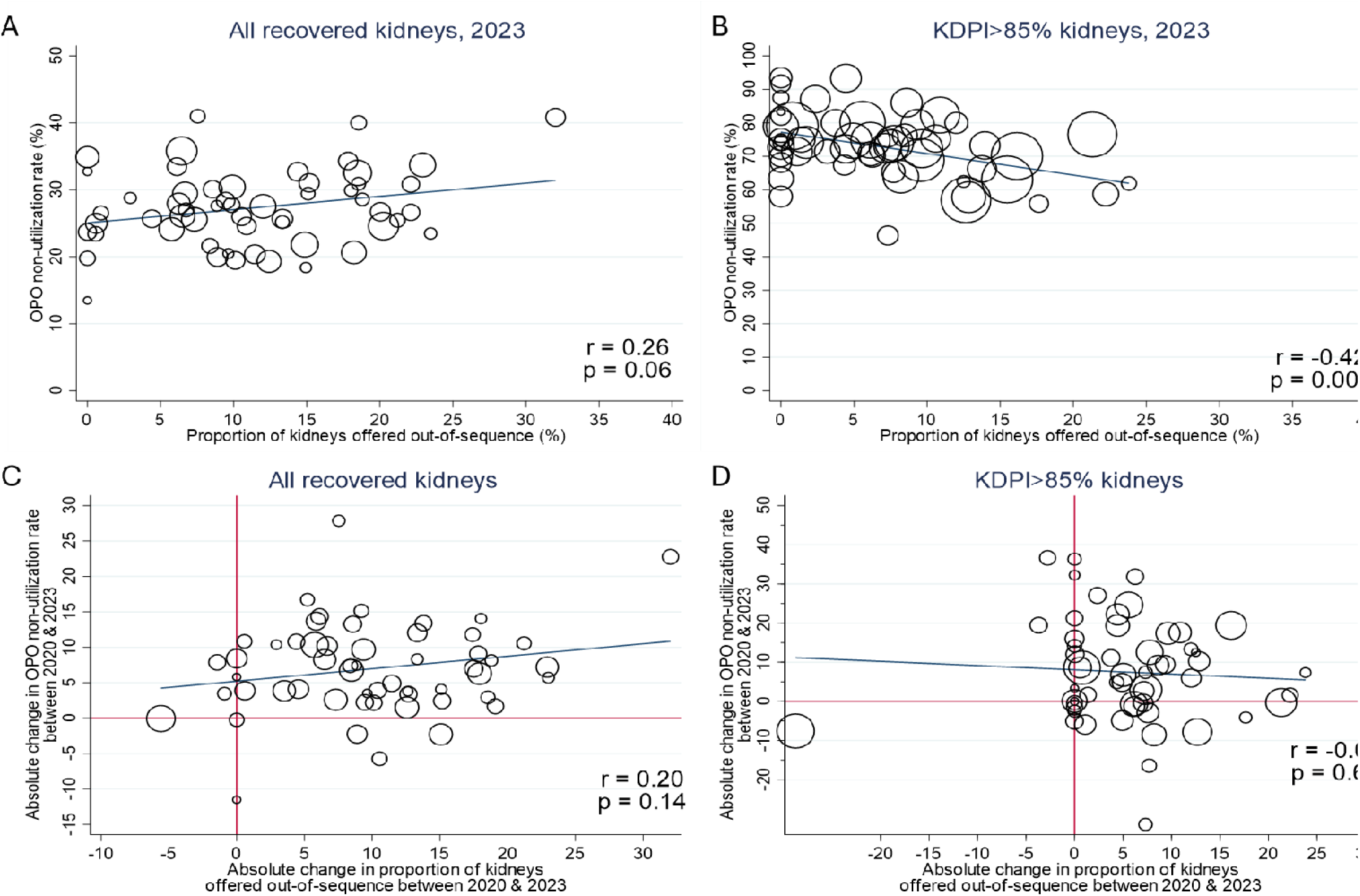
Correlations between the proportion of kidneys offered out-of-sequence by each OPO and their respective discard rates. Panels A and B show the association between the proportion of organs offered out-of-sequence by each of the OPOs in 2023 and the proportion of the procured deceased donor organs that are discarded for all recovered kidneys and those with a KDPI >85%, respectively. Panels C and D show the absolute change in the proportion of kidneys offered out-of-sequence and the absolute change in the proportion of kidneys discarded between 2020 and 2023. Each OPO is represented by a circle that is proportional in size to the number of deceased donor kidneys procured by that OPO.

### System-level distribution of out-of-sequence transplants

In 2020, there were relatively few transplant centers that received out-of-sequence deceased donor kidneys, with 53 centers (23% of US transplant centers) using at least 1 out-of-sequence deceased donor kidney and only 3 centers with >20% of their transplants resulting from out-of-sequence deceased donor kidneys (Supplemental Table 2). The extreme concentration in practice was reflected in the Gini coefficient of 0.93 (vs. 0.50 of in-sequence transplants) across centers (Figure 2). By 2023, the number of centers benefiting from out-of-sequence placements increased to 164 centers (71%) that used at least 1 out-of-sequence deceased donor kidney and 42 centers (18%) using out-of-sequence deceased donor kidneys for more than 20% of their transplants (Figure 4, Supplemental Figure 2). While more centers were receiving out-of-sequence deceased donor kidneys, the distribution of these transplants remained uneven, as evidenced by the Gini coefficient of 0.72 in 2023.

**Figure 4.**
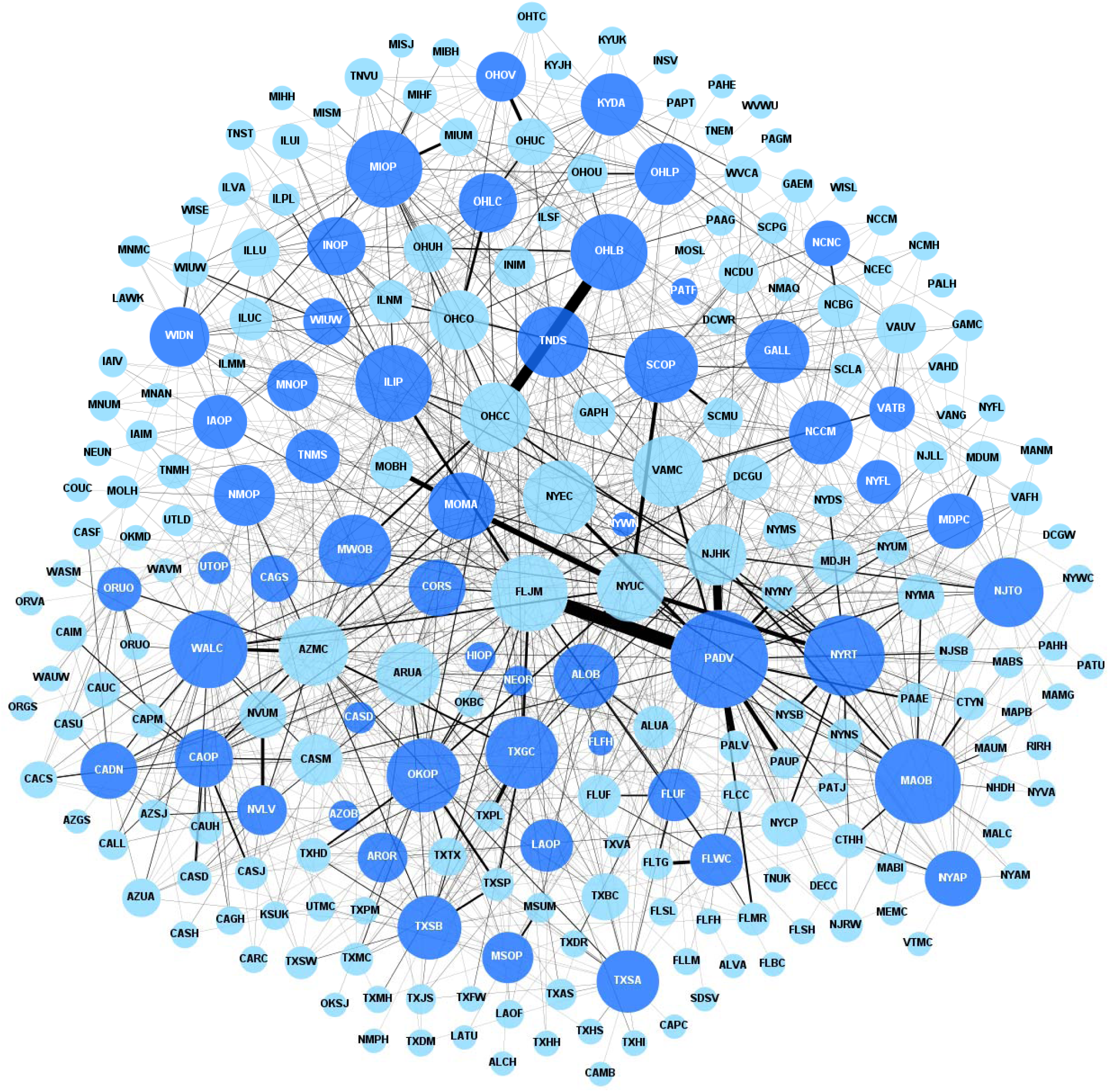
Network graph showing all out-of-sequence deceased donor transplant events between OPOs and transplant centers between 2020 and 2023. The thickness of the connecting lines is proportional to the number of events between the connected OPO and transplant center, and the size of the circle is proportional to the number of out-of-sequence transplants that the OPO or transplant center is participating in. OPOs are shown in dark blue and the transplant centers are shown in light blue.

Temporal trends in the Gini coefficient for out-of-sequence transplants were nearly identical for both high KDPI kidneys and lower KDPI kidneys (Supplemental Figure 3), suggesting that out-of-sequence allocation is not more selectively utilized by OPOs for high KDPI kidneys. In 2023, 27% of KDPI>85% deceased donor kidney transplants were out-of-sequence, while 15% of transplants with KDPI≤ 85% were also out of sequence (Supplemental Table 3).

### Recipients of kidneys placed out-of-sequence

Recipients of deceased donor kidneys allocated out-of-sequence were older, had a higher prevalence of diabetes, and included a higher proportion of non-Hispanic White males and individuals with private insurance (Table 1). These recipients also tended to have higher education attainment levels. Nearly double the proportion of recipients of out-of-sequence kidneys were pre-emptive compared to recipients of in-sequence allocation, while the recipients of out-of-sequence kidneys had a significantly shorter time on dialysis than the recipients of in- sequence kidneys.

**Table 1.** Comparison of recipient characteristics of deceased donor kidneys that were allocated.

|  | **In-sequence recipients**<br>**n = 61,497 (91.5%)** | **OOS recipients**<br>**n = 5,703 (8.5%)** | **p-value** |
| --- | --- | --- | --- |
| **Age at transplant** | 54 (42 - 63) | 61 (51 - 68) | <0.0001 |
| **Race** |  |  | <0.0001 |
| Black | 21,640 (35.2%) | 1,800 (31.5%) |  |
| White | 33,715 (54.8%) | 3,169 (55.5%) |  |
| Other/Unknown | 6,142 (10.0%) | 734 (13.0%) |  |
| **Ethnicity** |  |  | 0.007 |
| Hispanic | 12,712 (20.7%) | 1,093 (19.2%) |  |
| Non-Hispanic | 48,785 (79.3%) | 4,610 (80.8%) |  |
| **Gender** |  |  | <0.0001 |
| Female | 24,850 (40.4%) | 1,956 (34.3%) |  |
| Male | 36,647 (59.6%) | 3,747 (65.7%) |  |
| **History of diabetes** |  |  | <0.0001 |
| Yes | 22,508 (36.6%) | 2,610 (45.8%) |  |
| No/Unknown | 39,989 (63.4%) | 3,093 (54.2%) |  |
| **Education** |  |  | <0.0001 |
| None/Unknown | 1,501 (2.4%) | 122 (2.1%) |  |
| High school/GED or less | 28,864 (46.9%) | 2,322 (40.7%) |  |
| At least some college | 31,132 (50.7%) | 3,261 (57.2%) |  |
| **Primary payor at transplant** |  |  | <0.0001 |
| Public | 46,247 (75.2%) | 4,017 (70.4%) |  |
| Private | 15,239 (24.8%) | 1,685 (29.6%) |  |
| Other/Unknown | 11 (<1%) | 1 (<1%) |  |
| **CIT (hours)** | 18.5 (13.9 - 23.0) | 24.4 (20.3 - 29.9) | <0.0001 |
| **KDPI** | 39 (20 - 60) | 56 (33 - 73) | <0.0001 |
| **Preemptive transplant** | 6,749 (11.0%) | 1,051 (18.4%) | <0.0001 |
| **Dialysis time (years), if non-preemptive** | 4.5 (2.3 - 7.2) | 2.5 (1.4 - 4.1) | <0.0001 |

## Discussion

This analysis of U.S. nationwide transplant registry data demonstrates a marked increase in out-of-sequence allocation of deceased donor kidneys over time, with greater adoption of this practice by the majority of OPOs that have directed these out-of-sequence organs to a relatively select group of transplant centers. Notably, there is scant evidence that these efforts are associated with improved organ utilization patterns, despite their widespread adoption.

Out-of-sequence deceased donor kidney transplants have always existed but were rare and used primarily by just 3 OPOs.[14] This previously rare violation of allocation policy has now become a widespread practice, with out-of-sequence allocation utilized for a rapidly growing proportion of deceased donor kidneys.[20] Several OPOs and transplant centers use out-of-sequence allocation as the predominant pathway for allocation, resulting in the movement of organs preferentially to select transplant centers, thereby exacerbating inequities in access to transplantation for waitlisted patients.[21]

The transplant center-level Gini coefficient of 0.72 for out-of-sequence placements in 2023 highlights just how concentrated the allocation of out-of-sequence deceased donor kidneys is at present – despite the widespread participation of the OPOs, reflected by the decreasing Gini coefficient for OPOs from 0.89 in 2020 to 0.47 in 2023. This concentration of out-of-sequence transplants at select centers may exacerbate geographic disparities that the most recent allocation changes were designed to overcome. Perhaps even worse would be the eventual inadvertent creation of a 2-tiered transplant system, with some centers having preferential access to the supply of deceased donor kidneys compared to others. Access to transplantation for waitlisted patients is already highly variable between centers as a result of large differences in organ offer acceptance rates.[6 21] Allowing out-of-sequence placements to grow while the organ shortage intensifies risks further exaggerating differences in access to transplantation for patients waitlisted at different centers.[22 23]

Given the higher discard rate of high KDPI (i.e., lower-quality) kidneys, it is possible that out-of-sequence allocation is contributing to improved utilization of these organs. However, a significant portion of out-of-sequence deceased donor kidney transplants include some of the highest-quality organs that would otherwise be preferentially allocated to the healthiest recipients. The proportion of deceased donor kidneys placed out-of-sequence with a KDPI>85% was similar to those ≤85% and the Gini coefficient was similar for these strata of kidneys, suggesting that OPOs are not restricting the use of out-of-sequence allocation to only kidneys at high-risk of discard. This raises the concern that, without oversight, many deceased donor kidneys may have been inappropriately placed out-of-sequence to the detriment of the skipped over centers and their patients. This also raises important questions about when deceased donor kidneys should be offered through the out-of-sequence pathway and to which center. It is also unclear what objective criteria are used by centers accepting these offers to ensure that these organs are being allocated to waitlisted individuals fairly and equitably.

Additionally, the absence of an association between increasing proportions of out-of-sequence transplants and a subsequent reduction in the discard rate would suggest that this is not the panacea to allocation inefficiency and discards that it is perceived to be. Instead, out-of-sequence allocation appears to favor select centers and encourage centers to identify a patient by list diving – a practice that further erodes the objective nature of allocation policy.[24] While many less-than-ideal organs may avoid discard with an expedited placement pathway, this needs to be carefully designed, objective, and transparent, with clear definitions for which organs would qualify, and how centers could opt in.[25–27] The risk of exacerbating inequities that result from out-of-sequence organ placements and subsequent list-diving risks needs to be recognized and offset with greater transparency and potentially greater regulatory oversight.

Finally, OPOs favoring one transplant center over another also risks undermining efforts from the Center for Medicare and Medicaid Innovation to increase rates of kidney transplantation in the United States.

In conclusion, we demonstrate the increasing proportion of deceased donor kidneys being placed out-of-sequence by OPOs, with the majority placed at a relatively small set of transplant centers. This rapidly growing practice is not restricted to lower-quality deceased donor kidneys and appears to be exacerbating existing inequities in access to transplantation by creating preferential access for centers, encouraging list diving, and undermining the objective nature of the allocation system. The transplant system urgently needs a more efficient objective allocation system that strives for increased equity while facilitating the efficient placement of deceased donor kidneys. Subjective workarounds like out-of-sequence allocation will undermine public trust to the detriment of patients and the transplant system as a whole.

## Data Availability

The data that support the findings of this study are available by request from the Scientific Registry of Transplant Recipients (SRTR). Restrictions apply to the sharing of these data due to privacy or ethical restrictions.

## Supplemental Material

Supplemental Table 1. **Error! Bookmark not defined.**

*The absolute number of out-of-sequence deceased donor kidneys in each KDPI decile that were transplanted each year and the proportion of all deceased donor kidney transplants that were out-of-sequence in each KDPI decile*

Supplemental Table 2. **Error! Bookmark not defined.**

*Distribution of out-of-sequence transplants across transplant centers in the United States*.

Supplemental Table 3. **Error! Bookmark not defined.**

*The proportion of out-of-sequence deceased donor kidney transplants among all transplants in 2020-2023, stratified by high KDPI status*.

Supplemental Figure 1 **Error! Bookmark not defined.**

*STROBE diagram of study cohort*

Supplemental Figure 2 **Error! Bookmark not defined.**

*Network graph showing all in-sequence deceased donor transplant events between OPOs and transplant centers between 2020 and 2023*.

Supplemental Figure 3 **Error! Bookmark not defined.**

*Temporal trends in the proportion of out-of-sequence deceased donor kidney transplants*.

## References

1. Adler JT, Husain SA, King KL, Mohan S. Greater complexity and monitoring of the new Kidney Allocation System: Implications and unintended consequences of concentric circle kidney allocation on network complexity. American journal of transplantation : official journal of the American Society of Transplantation and the American Society of Transplant Surgeons 2021;21(6):2007–13 doi: 10.1111/ajt.16441 [published Online First: 20210102].

2. Israni AK, Salkowski N, Gustafson S, et al. New national allocation policy for deceased donor kidneys in the United States and possible effect on patient outcomes. Journal of the American Society of Nephrology: JASN 2014;25(8):1842.

3. Puttarajappa CM, Hariharan S, Zhang X, et al. Early Effect of the Circular Model of Kidney Allocation in the United States. Journal of the American Society of Nephrology : JASN 2022 doi: 10.1681/asn.2022040471 [published Online First: 2022/10/28].

4. Israni AK, Salkowski N, Gustafson S, et al. New national allocation policy for deceased donor kidneys in the United States and possible effect on patient outcomes. Journal of the American Society of Nephrology 2014;25(8):1842–48.

5. Husain SA, King KL, Pastan S, et al. Association Between Declined Offers of Deceased Donor Kidney Allograft and Outcomes in Kidney Transplant Candidates. JAMA network open 2019;2(8):e1910312–e12 doi: 10.1001/jamanetworkopen.2019.10312.

6. Mohan S, Chiles MC. Achieving equity through reducing variability in accepting deceased donor kidney offers. Clin J Am Soc Nephrol 2017;12(8):1212.

7. Concepcion BP, Harhay M, Ruterbories J, et al. Current landscape of kidney allocation: Organ procurement organization perspectives. Clin Transplant 2023;37(4):e14925 doi: 10.1111/ctr.14925 [published Online First: 20230212].

8. Cron DC, Husain SA, King KL, Mohan S, Adler JT. Increased volume of organ offers and decreased efficiency of kidney placement under circle-based kidney allocation. American journal of transplantation : official journal of the American Society of Transplantation and the American Society of Transplant Surgeons 2023;23(8):1209–20 doi: 10.1016/j.ajt.2023.05.005 [published Online First: 20230516].

9. Mohan S, Yu M, King KL, Husain SA. Increasing Discards as an Unintended Consequence of Recent Changes in United States Kidney Allocation Policy. Kidney Int Rep 2023;8(5):1109–11 doi: 10.1016/j.ekir.2023.02.1081 [published Online First: 20230225].

10. Mohan S, Yu M, King KL, Husain SA. Increasing discards as an unintended consequence of recent changes in United States kidney allocation policy. Kidney International Reports 2023;8(5):1109.

11. Reddy V, da Graca B, Martinez E, et al. Single-center analysis of organ offers and workload for liver and kidney allocation. American Journal of Transplantation 2022;**22**(11):2661-67.

12. Medicare and Medicaid Programs; Organ Procurement Organizations Conditions for Coverage: Revisions to the Outcome Measure Requirements for Organ Procurement Organizations. In: Services CfMM, ed. Federal Register, 2021:77898-949.

13. Goldberg D, Karp S, Shah MB, Dubay D, Lynch R. Importance of incorporating standardized, verifiable, objective metrics of organ procurement organization performance into discussions about organ allocation. American Journal of Transplantation 2019;19(11):2973–78.

14. King KL, Husain SA, Perotte A, Adler JT, Schold JD, Mohan S. Deceased donor kidneys allocated out of sequence by organ procurement organizations. American journal of transplantation : official journal of the American Society of Transplantation and the American Society of Transplant Surgeons 2022;22(5):1372–81 doi: 10.1111/ajt.16951 [published Online First: 2022/01/10].

15. King KL, Chaudhry SG, Ratner LE, Cohen DJ, Husain SA, Mohan S. Declined offers for deceased donor kidneys are not an independent reflection of organ quality. Kidney 360 2021;2(11):1807.

16. Lewis Z. OPTN Membership and Professional Standards Committee Report to the Board of Directors. Secondary OPTN Membership and Professional Standards Committee Report to the Board of Directors 2023. https://optn.transplant.hrsa.gov/media/lkunawmp/membership-and-professional-standards-committee-mpsc-report-to-the-board-june-2023.pdf.

17. Lorenz M. Methods of Measuring the Concentration of Wealth. Publications of the American Statistical Association 1905;9(70):209–19.

18. Regidor E. Measures of health inequalities: part 1. Journal of epidemiology and community health 2004;58(10):858.

19. Catalano MT, Leise TL, Pfaff TJ. Measuring resource inequality: The Gini coefficient. Numeracy 2009;2(2):4.

20. Wood N, GR L, JJ S. Deviating from the match run to save a kidney American journal of transplantation : official journal of the American Society of Transplantation and the American Society of Transplant Surgeons 2023.

21. King KL, Husain SA, Schold JD, et al. Major variation across local transplant centers in probability of kidney transplant for wait-listed patients. Journal of the American Society of Nephrology 2020;31(12):2900–11.

22. King KL, Husain SA, Cohen DJ, Schold JD, Mohan S. The role of bypass filters in deceased donor kidney allocation in the United States. American journal of transplantation : official journal of the American Society of Transplantation and the American Society of Transplant Surgeons 2022;22(6):1593–602 doi: 10.1111/ajt.16967 [published Online First: 2022/01/29].

23. Yu M, King KL, Husain SA, Schold JD, Mohan S. Use of Offer Bypass Filters Under the Updated Kidney Allocation System. Kidney 360 2024:10.34067.

24. King KL, Husain SA, Yu M, Adler JT, Schold J, Mohan S. Characterization of transplant center decisions to allocate kidneys to candidates with lower waiting list priority. JAMA network open 2023;6(6):e2316936–e36.

25. Jadlowiec CC, Ohara SY, Punukollu R, et al. Outcomes with transplanting kidneys offered through expedited allocation. Clin Transplant 2023;37(11):e15094 doi: 10.1111/ctr.15094 [published Online First: 20230810].

26. Kilambi V, Barah M, Formica RN, Friedewald JJ, Mehrotra S. Evaluation of Opening Offers Early for Deceased Donor Kidneys at Risk of Nonutilization. Clinical Journal of the American Society of Nephrology 2023:10.2215.

27. Noreen SM, Klassen D, Brown R, et al. Kidney accelerated placement project: Outcomes and lessons learned. American journal of transplantation : official journal of the American Society of Transplantation and the American Society of Transplant Surgeons 2022;22(1):210–21 doi: 10.1111/ajt.16859 [published Online First: 20211025].

