## Supplement for "Out-of-sequence placement of deceased donor kidneys is exacerbating inequities in the United States"

*Temporal trends in the proportion of out-of-sequence deceased donor kidney transplants.*

**Supplemental Table 1.** The proportion of all deceased donor kidney transplants that were out-of-sequence in each KDPI decile in each of the 4 years.

| Year | 0-9% | 10-19% | 20-29% | 30-39% | 40-49% | 50-59% | 60-69% | 70-79% | 80-89% | 90-100% |
|---|---|---|---|---|---|---|---|---|---|---|
| **2020** | 0.5% | 1.0% | 2.1% | 2.4% | 2.5% | 2.1% | 3.6% | 3.8% | 3.7% | 3.9% |
| **2021** | 1.9% | 2.9% | 4.9% | 3.8% | 4.3% | 4.7% | 6.9% | 7.4% | 4.9% | 6.9% |
| **2022** | 3.9% | 4.8% | 7.7% | 8.6% | 8.8% | 11.6% | 13.7% | 15.5% | 17.7% | 18.8% |
| **2023** | 6.9% | 8.7% | 10.7% | 13.8% | 14.2% | 16.9% | 21.2% | 25.0% | 25.4% | 30.9% |

**Supplemental Table 2.** Distribution of out-of-sequence transplants across transplant centers in the United States.

| | **2020** | **2021** | **2022** | **2023** |
|---|---|---|---|---|
| Total number of centers with at least 1 DDK transplant | 233 | 232 | 235 | 231 |
| Number of centers with no out-of-sequence DDK transplants (n) | 180 | 128 | 85 | 67 |
| Number of centers with at least 1 out-of-sequence DDK transplant (n) | 53 | 104 | 150 | 164 |
| Number of centers with ≤ 5 out-of-sequence DDK transplants (n) | 223 | 199 | 167 | 132 |
| Number of centers with > 5 out-of-sequence DDK transplants (n) | 10 | 33 | 68 | 99 |
| Number of centers with > 20% DDK transplants that are out-of-sequence (n) | 3 | 5 | 17 | 42 |
| Number of centers with > 50% DDK transplants that are out-of-sequence (n) | 0 | 0 | 3 | 4 |
| Median number of out-of-sequence DDKs at transplant center | 0 | 0 | 1 | 4 |
| Median proportion of out-of-sequence DDKs at transplant center (%) | 0% | 0% | 2.5% | 6.3% |
| Highest number of out-of-sequence DDKs at a single center (n) | 129 | 71 | 168 | 148 |
| Highest proportion of DDKs at a single center that are out-of-sequence (%) | 38.1% | 29.2% | 52.5% | 62.7% |

**Supplemental Table 3.** The proportion of out-of-sequence deceased donor kidney transplants among all transplants in 2020-2023, stratified by high KDPI status.

| | Year | In-sequence Gini | Out-of-sequence Gini | Total deceased donor kidney transplants | Out-of-sequence Transplants | Proportion out-of-sequence transplants |
|---|---|---|---|---|---|---|
| **KDPI ≤ 85%** | **2020** | 0.50 | 0.94 | 14,338 | 309 | 2.2% |
| | **2021** | 0.50 | 0.81 | 15,547 | 696 | 4.5% |
| | **2022** | 0.48 | 0.78 | 16,410 | 1,537 | 9.4% |
| | **2023** | 0.47 | 0.72 | 17,464 | 2,623 | 15.0% |
| **KDPI > 85%** | **2020** | 0.50 | 0.96 | 675 | 31 | 4.6% |
| | **2021** | 0.50 | 0.91 | 769 | 46 | 6.0% |
| | **2022** | 0.49 | 0.80 | 863 | 153 | 17.7% |
| | **2023** | 0.50 | 0.72 | 1,134 | 308 | 27.2% |

**Supplemental Figure 1.** STROBE diagram of study cohort.

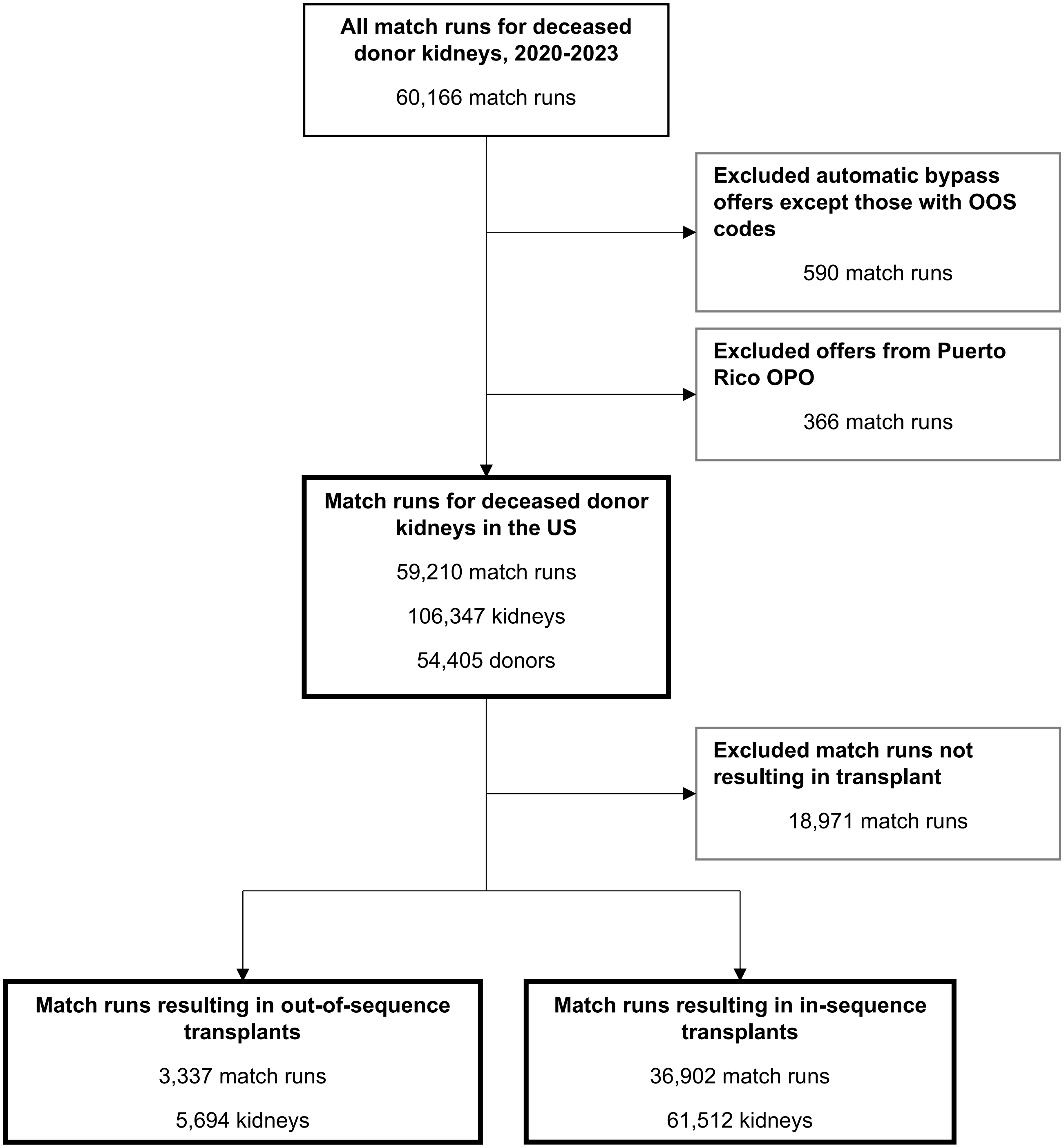

**Supplemental Figure 2.** Network graph showing all in-sequence deceased donor transplant events between OPOs and transplant centers between 2020 and 2023. The thickness of the connecting lines is proportional to the number of events between the connected OPO and transplant center, and the size of the circle is proportional to the number of in-sequence transplants that the OPO or transplant center is participating in. OPOs are shown in dark blue and the transplant centers are shown in light blue.

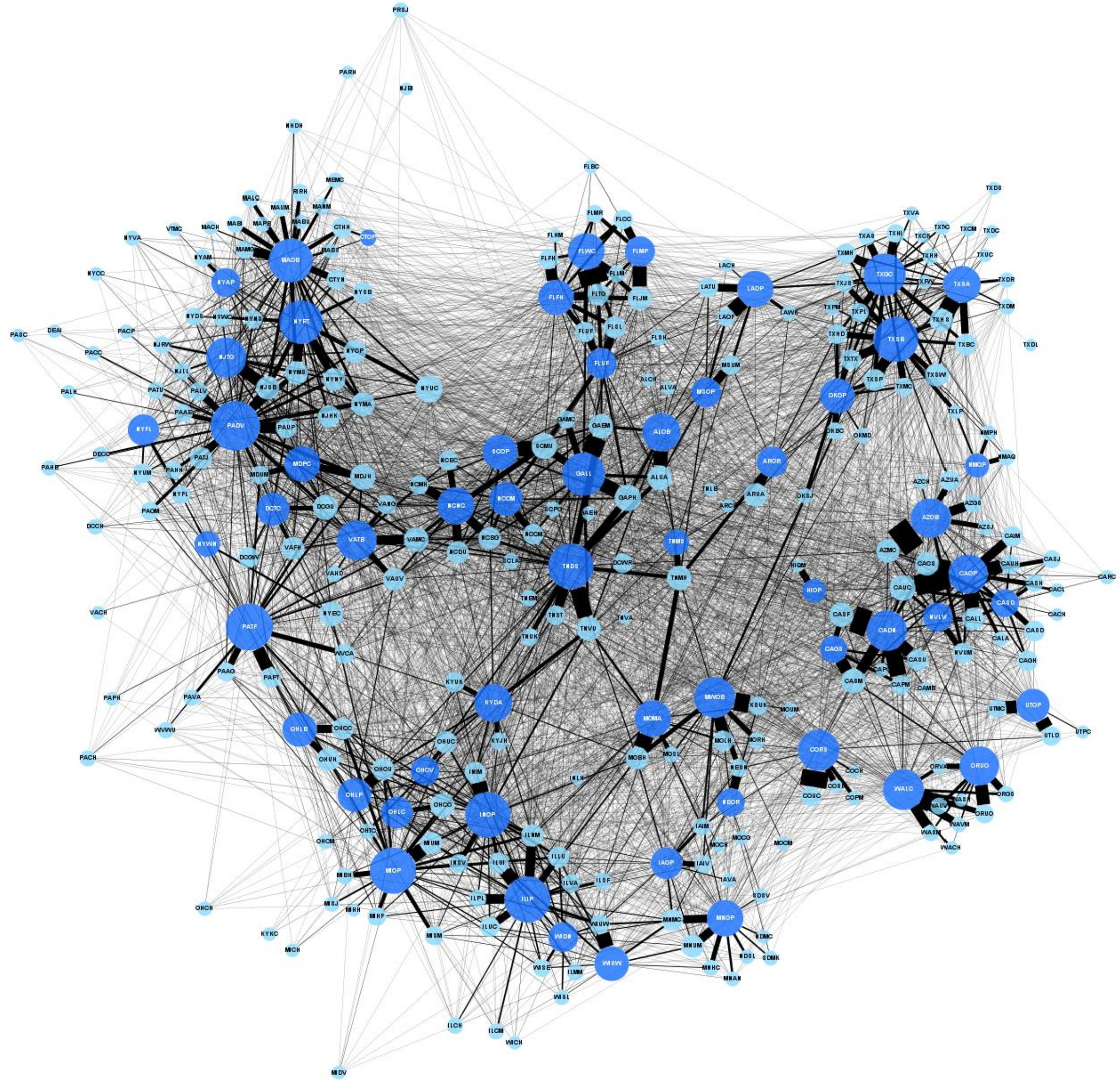

**Supplemental Figure 3.** Temporal trends in the proportion of out-of-sequence deceased donor kidney transplants. The red lines connect the Gini coefficient for out-of-sequence transplants and green lines connect the Gini coefficient for in-sequence transplants. The solid lines connect the Gini coefficients for KDPI ≤ 85% and dashed lines for KDPI >85%.

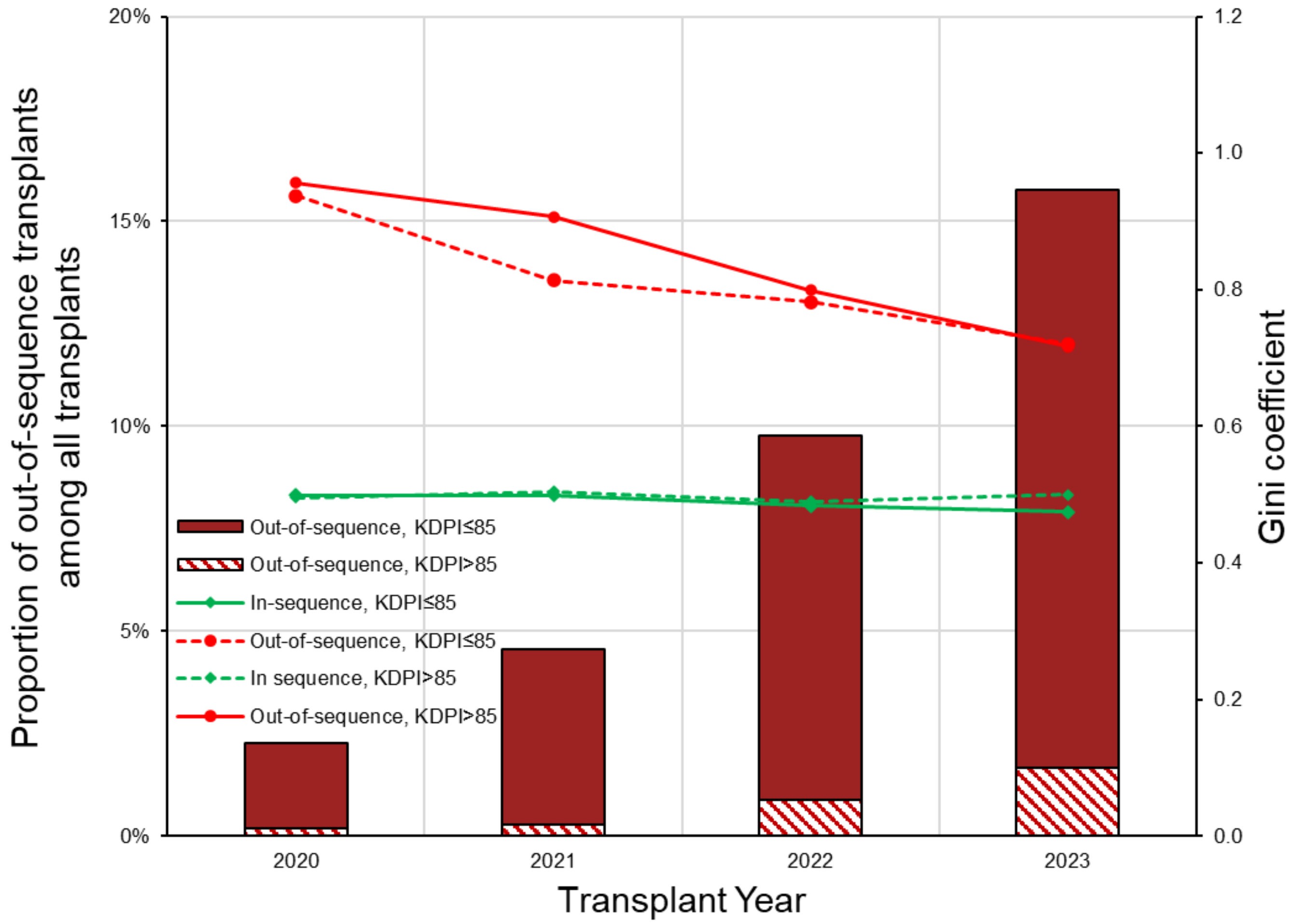
